# Belief of Previous COVID-19 Infection and Unclear Government Policy are Associated with Reduced Willingness to Participate in App-Based Contact Tracing: A UK-Wide Observational Study of 13,000 Patients

**DOI:** 10.1101/2020.06.03.20120337

**Authors:** Patrik Bachtiger, Alexander Adamson, Jennifer K Quint, Nicholas S Peters

## Abstract

**Background:** Contact tracing and lockdown are health policies being used worldwide to combat the coronavirus (COVID-19). While easing lockdown, the UK National Health Service (NHS) launched its Track and Trace Service at the end of May 2020, and aims by end of June 2020 also to include app-based notification and advice to self-isolate for those who have been in contact with a person known to have COVID-19. To be successful, such an app will require high uptake, the determinants and willingness for which are unclear but essential to understand for effective public health benefit.

**Objectives:** To measure the determinants of willingness to participate in an NHS app-based contact tracing programme using a questionnaire within the Care Information Exchange (CIE) - the largest patient-facing electronic health record in the NHS.

**Methods:** Observational study of 47,708 registered NHS users of the CIE, 27% of whom completed a novel questionnaire asking about willingness to participate in app-based contact tracing, understanding of government advice, mental and physical wellbeing and their healthcare utilisation -- related or not to COVID-19. Descriptive statistics are reported alongside univariate and multivariable logistic regression models, with positive or negative responses to a question on app-based contact tracing as the dependent variable.

**Results:** 26.1% of all CIE participants were included in the analysis (N = 12,434, 43.0% male, mean age 55.2). 60.3% of respondents were willing to participate in app-based contact tracing. Out of those who responded ‘no’, 67.2% stated that this was due to privacy concerns. In univariate analysis, worsening mood, fear and anxiety in relation to changes in government rules around lockdown were associated with lower willingness to participate. Multivariable analysis showed that difficulty understanding government rules was associated with a decreased inclination to download the app, with those scoring 1-2 and 3-4 in their understanding of the new government rules being 45% and 27% less inclined to download the contact tracing app, respectively; when compared to those who rated their understanding as 5-6/10 (OR for 1-2/10 = 0.57 [CI 0.48 - 0.67]; OR for 3-4/10 = 0.744 [CI 0.64 - 0.87]), whereas scores of 7-8 and 9-10 showed a 43% and 31% respective increase. Those reporting an unconfirmed belief of having previously had and recovered from COVID-19 were 27% less likely to be willing to download the app; belief of previous recovery from COVID-19 infection OR 0.727 [0.585 - 0.908]).

**Conclusions:** In this large UK-wide questionnaire of wellbeing in lockdown, a willingness for app-based contact tracing is 60% - close to the estimated 56% population uptake, and substantially less than the smartphone-user uptake considered necessary for an app-based contact-tracing to be an effective intervention to help suppress an epidemic. Given this marginal level of support over an appropriate age range, the impacts of difficulty comprehending government advice and a policy of not testing to confirm self-reported COVID-19 infection during lockdown indicate that uncertainty in communication and diagnosis in adopted public health policies will negatively impact the effectiveness of a government contact tracing app.

## INTRODUCTION

Coronavirus disease 2019 (COVID-19), caused by severe acute respiratory syndrome–coronavirus 2 (SARS-CoV-2), has acutely incapacitated health systems, but there is growing concern that COVID-19 will be a long-lasting and fluctuating pandemic.^1-3^ In the absence of an effective vaccine, so-called non-pharmacological interventions have become the vanguard for reducing viral transmission by suppressing contact rates in the population.^4,5^ These include physical distancing, decontamination and hygiene measures -- as well as case identification and isolation with contact tracing and quarantine.

The UK government launched its COVID-19 contact tracing programme, the National Health Service (NHS) Test and Trace Service, at the end of May 2020.^6^ In the lead up to this announcement, NHSX, the unit responsible for setting national policy for use of technology in the NHS, had led on the piloting of an app on the Isle of Wight, with plans to later operationalise this nationally as part the wider contact tracing strategy (by the end of June 2020). Individuals would be asked to use their mobile phone to download the app, which will run in the background using low-energy Bluetooth technology to notify users if they have been in close proximity for more than 15 minutes with someone known to be COVID-19 positive, while maintaining anonymity. They would subsequently be advised to self-isolate at home for 14 days.

Germany,^7^ South Korea,^8^ Singapore^9^ and Hong Kong^10^ have all deployed extensive testing and contact tracing -- notably much earlier than the UK -- and so far have experienced comparatively lower fatality rates.^11^ A phone app is cheap, scalable and could show how finite resources maybe divided between different intervention strategies for the most effective control.^12,13^

To date there is inadequate understanding of the public’s willingness to participate in an app for contact tracing for COVID-19, and how the context of existing public health policy may influence this willingness. Part of the challenge is inadequate sampling from those with health conditions - - i.e. patients -- who as an at-risk group^14,15^ stand to benefit most from participation in app-based contact tracing. An online poll in May 2020 by Opinium surveyed 2,002 participants, with half (53%) stating they would be likely to download a contact tracing app, while 21% would be unlikely to.^16^ A further, non-representative poll of 730 participants suggested 73% were willing to download an app.^17^

The exact proportion of engagement required for effectiveness is not yet known; the only modelling thus far addressing this question comes from a report submitted to NHSX by the Oxford University Big Data institute, suggesting effective epidemic suppression with 80% of all smartphone users using the app, or 56% of the population overall.^18^ However, the potential determinants of this willingness to participate, such as age, sex, healthcare service utilisation, impact of changing government rules on lockdown on wellbeing and perceptions of own experience of COVID-19, remain unknown.

## METHODS

### Study Participants

Participants in this study were individuals with a previous healthcare event or encounter e.g. hospital admission, outpatient appointment, medical investigation at a London-based tertiary NHS Trust (Imperial College Healthcare NHS Foundation Trust). Any such encounter triggered the creation of a digital record on the NHS Trust’s Care Information Exchange (CIE)^19^, the UK’s largest patient-facing electronic health record (EHR), displaying, for example, appointments, clinical letters, blood results, accessible to patients after registration with an email address. On the date on which the data were extracted (27th May 2020) the CIE held the records of 47,679 patients aged 18 years or older.

### Questionnaire Design

The CIE has several functionalities for enhancing direct patient care, including the ability to create bespoke questionnaires. Invitation to respond to questionnaires is notified by email, with a direct web link for completion within a patient’s CIE record. The data analysed in this study were derived from a single questionnaire that was part of a longitudinal, weekly series implemented at the beginning of lockdown as a direct care tool for patients to keep track of their wellbeing.

### Timing of Questionnaire

The questionnaire was sent out on Friday 15th May 2020, five days after the UK government changed its messaging around lockdown from ‘Stay Home, Protect the NHS, Save Lives’ to ‘Stay Alert, Control the Virus, Save Lives’, accompanied by the easing of some lockdown restrictions.^20^ Responses submitted after Monday 18th May 2020 were excluded to minimise recall bias and ensure questions referring to ‘in the last week’ were not misinterpreted and were pertinent to the events of that week.

### Questionnaire content

The questionnaire used for this study included the addition of a question to measure a participant’s willingness to participate in app-based contact tracing, alongside established questions that formed part of the longitudinal weekly questionnaires for patients to track their wellbeing. Data relevant to analysis were: age, sex, understanding of changing government rules on lockdown; effect of changing government rules on lockdown on mood, anxiety and fear; experience of COVID-19 symptoms in last week, belief of previous COVID-19 illness and recovery, testing status for COVID-19, and healthcare contact since the start of lockdown. The full questionnaire is included in appendix A.

### Data and Consent

The CIE, including the questionnaire, is a tool for direct care at Imperial College Healthcare NHS Trust, therefore formal consent for data analysis and ethical approval was not required. Participants were informed that their responses would be analysed to help inform local and national health policy and were free to opt out. For the purposes of analysis with non-clinical collaborators, all data were de-identified in line with local information governance protocols, with the subsequent dataset approved for sharing by the Imperial College Healthcare NHS Trust Data Protection Office.

### Data analysis

All data were analysed using R (version 3.6.2). Questions with a response in the form of a five-point scale (a lot worse, a little worse, no change, a little better, a lot better) had their responses simplified to a three point scale (worse, no change, better) to aid interpretation of the results and account for low numbers in some categories. Age was categorised into 10 year age bands between 18-29 and 80+ to allow for a non-linear relationship between age and response to contact tracing. Participants who marked any one of the available options for healthcare encounters were classed as having any healthcare contact. Participants who marked themselves as having a new or worsened cough, a fever that was measured or unmeasured with a thermometer, or anosmia, were classed as displaying COVID-19 symptoms. Participants who stated that they had received any test for COVID-19 (positive/pending/negative) were classed as having received a test for COVID-19. Participants who stated that they had not received a test but thought that they had recovered from COVID-19, and additionally stated that they had received a test, were classed as having had a test and removed from the ‘no test but think recovered’ group (N = 8). Descriptive statistics are reported for the dataset as a whole and broken down according to response to inclination to download an NHS contact-tracing app (yes/no/unsure). Differences between groups were assessed using Chi-squared tests for categorical variables and analysis of variance tests for continuous variables. P values <0.05 were considered statistically significant. Relationships between inclination to download the app (yes vs. no) and each variable of interest were then assessed using univariate and multivariable logistic regression, with unadjusted odds ratios and odds ratios adjusted for every other variable in the model provided alongside 95% confidence intervals. Due to low numbers reporting testing positive or awaiting their test result, only the binary variable of receiving a test result yes/no was included in the multivariable analysis. Multi-collinearity was assessed by calculation of the variance inflation factor (VIF), with variables with a VIF >5 (indicating substantial multicollinearity) removed from the model.

## RESULTS

After excluding those aged <18, 13,095/47,679 (27.5%) individual responses were recorded, of which 12,452 (26.1%) were within the predetermined time frame (consort diagram appendix B). A further 18 participants did not answer the contact tracing question and were excluded, leaving a total of 12,434 (26.1%) patients included in the analysis. Compared to excluded and non-responders, timely respondents included a larger proportion of males (43.0% vs 36.6%, p<0.001) and were older on average (mean age 55.2 (SD 15.0) vs mean age 45.0 (SD 15.2), p < 0.001). A map of where CIE registrants live according to the first three letters of their postcode is included in appendix C, highlighting that this is a UK-wide population. Summary Table 1 reports baseline characteristics and associations between responses to the question on contact tracing and other measured variables.

**Table 1:** General characteristics and questionnaire responses of the study population. Table presents N and percentage of each category unless otherwise indicated. Variables are presented as a total and broken down according to patients’ responses to contact-tracing app. p-value for categorical variables represents the chi-squared test for difference between groups, and for continuous variables represents a one-way analysis of variance test.

| Variable |  | Total (N = 12434) | Yes (N = 7503, 60.3%) | No (N = 2132, 17.1%) | Not sure (N = 2799, 22.5%) | p |
| --- | --- | --- | --- | --- | --- | --- |
| Age | Mean (SD) | 55.2 (15.0) | 55.1 (15.0) | 55.6 (15.7) | 55.0 (14.7) | 0.321 |
| Gender | Male | 5346 (43.0) | 3263 (43.5) | 970 (45.5) | 1113 (39.8) | <0.001 |
|  | Female | 7088 (57.0) | 4240 (56.5) | 1162 (54.5) | 1686 (60.2) |  |
| Ease of understanding of new government rules; 1 = very difficult, 10 = very easy | 1-2 | 1494 (12.0) | 753 (10.0) | 390 (18.3) | 351 (12.5) | <0.001 |
|  | 3-4 | 2238 (18.0) | 1221 (16.3) | 472 (22.1) | 545 (19.5) |  |
|  | 5-6 | 2856 (23.0) | 1667 (22.2) | 471 (22.1) | 718 (25.7) |  |
|  | 7-8 | 3347 (26.9) | 2192 (29.2) | 434 (20.4) | 721 (25.8) |  |
|  | 9-10 | 2454 (19.7) | 1650 (22.0) | 355 (16.7) | 449 (16.0) |  |
|  | Missing | 45 (0.4) | 20 (0.3) | 10 (0.5) | 15 (0.5) |  |
| Effect of new government rules on mood | Better | 1737 (14.0) | 1205 (16.1) | 226 (10.6) | 306 (10.9) | <0.001 |
|  | No change | 6722 (54.1) | 4098 (54.6) | 1092 (51.2) | 1532 (54.7) |  |
|  | Worse | 3917 (31.5) | 2163 (28.8) | 802 (37.6) | 952 (34.0) |  |
|  | Missing | 58 (0.5) | 37 (0.5) | 12 (0.6) | 9 (0.3) |  |
| **Effect of new government rules on anxiety** | Better | 793 (6.4) | 546 (7.3) | 90 (4.2) | 157 (5.6) | <0.001 |
|  | No change | 7018 (56.4) | 4336 (57.8) | 1136 (53.3) | 1546 (55.2) |  |
|  | Worse | 4590 (36.9) | 2601 (34.7) | 896 (42.0) | 1093 (39.0) |  |
|  | Missing | 33 (0.3) | 20 (0.3) | 10 (0.5) | 3 (0.1) |  |
| **Effect of new government rules on fear** | Better | 993 (8.0) | 679 (9.0) | 118 (5.5) | 196 (7.0) | <0.001 |
|  | No change | 6584 (53.0) | 4036 (53.8) | 1110 (52.1) | 1438 (51.4) |  |
|  | Worse | 4821 (38.8) | 2771 (36.9) | 894 (41.9) | 1156 (41.3) |  |
|  | Missing | 36 (0.3) | 17 (0.2) | 10 (0.5) | 9 (0.3) |  |
| **Displaying COVID-19 symptoms in past week which require 7 days isolation according to the NHS** | Yes | 999 (8.0) | 598 (8.0) | 185 (8.7) | 216 (7.7) | 0.446 |
|  | No | 11435 (92.0) | 6905 (92.0) | 1947 (91.3) | 2583 (92.3) |  |
| **Patient has tested positive for COVID-19** | Yes | 57 (0.5) | 31 (0.4) | 14 (0.7) | 12 (0.4) | 0.328 |
|  | No | 12377 (99.5) | 7472 (99.6) | 2118 (99.3) | 2787 (99.6) |  |
| **Patient is awaiting a test result for COVID-19** | Yes | 76 (0.6) | 52 (0.7) | 10 (0.5) | 14 (0.5) | 0.349 |
|  | No | 12358 (99.4) | 7451 (99.3) | 2122 (99.5) | 2785 (99.5) |  |
| **Patient has tested negative for COVID-19** | Yes | 463 (3.7) | 294 (3.9) | 76 (3.6) | 93 (3.3) | 0.333 |
|  | No | 11971 (96.3) | 7209 (96.1) | 2056 (96.4) | 2706 (96.7) |  |
| **Patient has taken a test for COVID-19\*** | Yes | 588 (4.7) | 372 (5.0) | 98 (4.6) | 118 (4.2) | 0.274 |
|  | No | 11846 (95.3) | 7131 (95.0) | 2034 (95.4) | 2681 (95.8) |  |
| **Patient has not taken a test for COVID-19, but thinks that they have had it and recovered** | Yes | 600 (4.8) | 325 (4.3) | 124 (5.8) | 151 (5.4) | 0.005 |
|  | No | 11834 (95.2) | 7178 (95.7) | 2008 (94.2) | 2648 (94.6) |  |
| **Patient has received any healthcare contact since the start of lockdown** | Yes | 6494 (52.2) | 3898 (52.0) | 1105 (51.8) | 1491 (53.3) | 0.454 |
|  | No | 5940 (47.8) | 3605 (48.0) | 1027 (48.2) | 1308 (46.7) |  |

\*8 participants took more than one test for COVID-19 therefore the total N for this question is less than the total N of positive/pending/negative combined.

### Overall Willingness, Privacy Concerns and Responses by Sex and Age

Overall, 60.3% of participants responded ‘yes’ to being willing to participate in app-based contact tracing, with 17.1% responding ‘no’ and 22.5% responding that they were unsure. Out of those who responded ‘no’, 67.2% stated that this was due to privacy concerns, 21.9% stated that they did not have a smartphone or appropriate device, and 10.9% stated that they did not feel able to download the app.

Responses for yes and no did not differ significantly by sex, although females were more likely to state ‘not sure’ than males. Responses were similar in all age groups from ages 18-79, however those aged 80 and above were less likely to respond ‘yes’ to downloading the app when compared to younger respondents and were more likely to report being unable to download an app or not having a suitable mobile device (Table 2). Not being willing to participate on the grounds of privacy concerns was inversely associated with age.

### Univariate Logistic Regression

The results of the univariate logistic regression models are shown in Table 3. There was no association between age and a willingness to participate except in those aged above 80 (OR 0.50 [CI 0.36 - 0.70]). Likewise sex, being tested for COVID-19, receiving a positive or negative test result or awaiting reulst, and reporting COVID-19 symptoms (cough, fever, anosmia) were not significantly associated with a willingness to participate. If new government rules led to a worsening mood, anxiety or fear participants were less likely to respond ‘yes’ to willingness to download an app for contact tracing (OR 0.72 [CI 0.65 - 0.80]; OR 0.76 [CI 0.69 - 0.84]); OR 0.85 [CI 0.77 - 0.94]). A low understanding of government advice was associated with less willingness to download the app (understanding reported 1-2/10 vs 5-6; OR 0.55 [0.47 - 0.64]).

### Multivariable Logistic Regression

The results of the multivariable logistic regression are shown in Table 3. Multivariable analysis showed that difficulty in understanding government rules around lockdown was strongly associated with being less willing to download the app (understanding 1-2/10 OR 0.57 [CI 0.48 - 0.67]; understanding 3-4/10 OR 0.744 [CI 0.64 - 0.87]). Those who indicated that they found it easier to understand government advice were more likely to indicate that they would download the app (7-8/10 vs 5-6 OR 1.37 [CI1.18 - 1.59], 9-10/10 vs 5-6 OR 1.24 [1.06 to 1.46]). Belief of having previously had and recovered from COVID-19 was associated with being 28% less likely to be willing to participate (OR 0.72 [0.59 - 0.91]) in app based contact tracing (Table 3). In the multivariable analysis, being female was borderline associated with being willing to participate in contact tracing after adjusting for the effect of every other variable in the model. There was moderate multi-collinearity between changes in the effect of the new government rules on mood, anxiety, and fear (VIF range 1.59 to 2.27).

**Table 2:** Breakdown of responses to app-based contact tracing by age-group. P value represents chi-squared test for trend for age.

|  | Response to app based contact tracing |  |  |  |  |  |
| --- | --- | --- | --- | --- | --- | --- |
| Age cat. | "No - I do not feel able to do this" | "No - I do not have a smartphone / appropriate device" | "No - I have privacy concerns" | "Not sure" | "Yes" | Total |
| 18-29 | 5 (0.8) | 6 (1.0) | 100 (16.4) | 129 (21.1) | 370 (60.7) | 610 |
| 30-39 | 20 (1.1) | 12 (0.7) | 283 (15.6) | 401 (22.1) | 1097 (60.5) | 1813 |
| 40-49 | 39 (2.1) | 18 (1.0) | 262 (13.8) | 444 (23.5) | 1129 (59.7) | 1892 |
| 50-59 | 41 (1.5) | 54 (2.0) | 360 (13.0) | 647 (23.4) | 1658 (60.1) | 2760 |
| 60-69 | 63 (2.1) | 134 (4.5) | 291 (9.8) | 678 (22.7) | 1816 (60.9) | 2982 |
| 70-79 | 46 (2.3) | 164 (8.2) | 123 (6.2) | 432 (21.6) | 1231 (61.7) | 1996 |
| 80+ | 18 (4.7) | 79 (20.7) | 14 (3.7) | 68 (17.8) | 202 (53.0) | 381 |
| P-value | <0.001 | <0.001 | <0.001 | 0.398 | 0.876 |  |

**Table 3:** Associations between each variable of interest and inclination to download a contact tracing app (Yes vs No). Table presents results for univariate logistic regression analyses and multivariable logistic regression adjusted for every other variable in the model.

| Variable | Univariate analysis (odds ratios with 95% CI) | Multivariable analysis (odds ratios with 95% CI)* |
| --- | --- | --- |
| **Age (reference: 18-29)** |  |  |
| 30-39 | 1.04 (0.81 to 1.33) | 1.07 (0.83 to 1.38) |
| 40-49 | 1.06 (0.83 to 1.35) | 1.08 (0.84 to 1.38) |
| 50-59 | 1.09 (0.86 to 1.38) | 1.10 (0.86 to 1.40) |
| 60-69 | 1.12 (0.88 to 1.41) | 1.05 (0.82 to 1.33) |
| 70-79 | 1.11 (0.87 to 1.41) | 1.02 (0.79 to 1.30) |
| 80+ | 0.55 (0.40 to 0.75) | 0.50 (0.36 to 0.70) |
| Female | 1.08 (0.98 to 1.19) | 1.11 (1.00 to 1.24) |
| **Ease of understanding of new government rules (reference: 5-6)** |  |  |
| 1-2 | 0.55 (0.47 to 0.64) | 0.56 (0.48 to 0.66) |
| 3-4 | 0.73 (0.63 to 0.85) | 0.74 (0.64 to 0.86) |
| 7-8 | 1.43 (1.23 to 1.65) | 1.37 (1.18 to 1.59) |
| 9-10 | 1.31 (1.13 to 1.53) | 1.24 (1.06 to 1.46) |
| **Effect of new government rules on mood (reference: No change)** |  |  |
| Worse | 0.72 (0.65 to 0.80) | 0.90 (0.79 to 1.03) |
| Better | 1.42 (1.22 to 1.67) | 1.16 (0.97 to 1.38) |
| **Effect of new government rules on anxiety (reference: No change)** |  |  |
| Worse | 0.76 (0.69 to 0.84) | 0.97 (0.84 to 1.11) |
| Better | 1.59 (1.27 to 2.02) | 1.24 (0.96 to 1.62) |
| **Effect of new government rules on fear (reference: No change)** |  |  |
| Worse | 0.85 (0.77 to 0.94) | 1.07 (0.95 to 1.21) |
| **Better** | 1.58 (1.29 to 1.95) | 1.26 (1.02 to 1.58) |
| **Displaying covid-19 symptoms in past week which require 7 days isolation according to the NHS** | 0.91 (0.77 to 1.09) | 1.00 (0.83 to 1.19) |
| **Patient has tested positive for COVID-19** | 0.63 (0.34 to 1.22) | - |
| **Patient is pending a test result positive for COVID-19** | 1.48 (0.79 to 3.10) | - |
| **Patient has tested negative for COVID-19** | 1.10 (0.86 to 1.44) | - |
| **Patient has taken a test for COVID-19** | 1.08 (0.87 to 1.37) | 1.08 (0.86 to 1.38) |
| **Patient has not taken a test for COVID-19, but thinks that they have had it and recovered** | 0.73 (0.59 to 0.91) | 0.73 (0.59 to 0.91) |
| **Patient has received any healthcare contact since the start of lockdown** | 1.00 (0.91 to 1.11) | 1.03 (0.93 to 1.14) |

Multivariable analysis performed on 9512 patients. Those who answered unsure (N = 2799) or were missing data for any other variable (N = 123) were not included in the analysis. Univariate analysis performed with the variable of interest as the only predictor in the model. Multivariable analysis adjusted for every other variable in the model. Only patients receiving a test yes/no were included in the model due to low numbers in the groups testing positive and awaiting a test result.

## DISCUSSION

This study reports questionnaire responses from 12,434 participants from across the UK, measuring determinants of willingness to participate in the anticipated NHS app for COVID-19 contact tracing. Overall, 60.3% of respondents were willing to participate in app-based contact tracing, with 22.5% unsure. Among participants answering ‘no’, 67.2% stated that this was due to privacy concerns. Worsening mood, fear and anxiety were associated with reduced willingness to participate in app based contact tracing only by univariate analysis (likely due to moderate correlation between these variables by multivariable analysis). Multivariable analysis showed that difficulty in understanding government rules was associated with a decreased inclination to download the app, with those scoring 1-2 and 3-4 in their understanding of the new government rules being 45% and 27% less inclined to download the contact tracing app, respectively, whereas scores of 7-8 and 9-10 showed a 43% and 31% respective increase. Those reporting an unconfirmed belief of having previously had and recovered from COVID-19 were 27% less likely to be willing to download the app.

This is the first instance of using the questionnaire functionality across the NHS’ largest patient-facing electronic health record (CIE) to collect, at scale, responses from a group of patients from across the UK. Other, conventional online questionnaires/polling platforms are characterised by selection bias, unknown denominators, inherently more digitally literate participants^21^ and limited external validity.^22^ This study therefore also highlights the opportunities for the CIE as a tool for population health surveillance, both in the short-term of the COVID-19 pandemic and as the NHS continues with its digital transformation agenda.^23^

The principal finding of this study that overall 60% are willing to participate in app-based contact tracing is close to the estimated 56% of the total population, but far less than the estimated 80% of smartphone users, needed for the app to have beneficial impact on an epidemic.^18^

The timing of the questionnaire coincided with changes in government announcements of lockdown policy which for many created substantial uncertainty, and this might be expected to reduce motivation, and possibly trust, as an explanation for the observed association between unwillingness for app based contact tracing and difficulty understanding government advice.

The UK adopted a public health policy during lockdown of instruction to stay at home if symptomatic of COVID-19 unless becoming very unwell with it,^11,24,25^ resulting in a large number of people who believe they have had COVID-19 but without confirmatory testing. Participants in this questionnaire who reported the unproven belief of having had COVID-19 may be less willing to participate on account of believing they may have immunity. Feeling the individual threat of COVID-19 no longer applies to them, this may also diminish the incentive to participate in the app to avoid risking being asked to potentially self-isolate for 14 days, at present regardless of perceived or objective history of COVID-19.

Use of CIE and participation in this study arguably implies a good level of digital literacy, however the age distribution of the sample is not skewed towards younger participants. Importantly, the ‘digital divide’, which suggests that those in their seventies are unable to participate in health initiatives with a digital element,^26^ is not reflected in this study.

There are several limitations to this study. These results are only indicative; whether participants stick to their response when faced with wide release and accompanying messaging from the government to download the app is uncertain. However, previous studies have shown good correlation between declared survey responses and subsequent behaviour.^27-29^ Twenty-two percent of respondents were ‘not sure’ about their willingness, but this response was not further qualified to identify underlying reasons. Patients registered on the CIE without internet, who could not log in to their CIE account, or were incapable of understanding or responding will be underrepresented, though such potential biases will have been mitigated by the large sample size. This study will by default have included many ‘shielded’ patients, identified and advised by the NHS to stay at home at all times due to their disease profile placing them at higher risk of adverse outcomes from COVID-19,^16^ for whom attitudes to participation in app based contact tracing will have its own considerations but are no less important.

### Conclusion

Poor understanding of government rules around lockdown and belief of having had COVID-19 decrease willingness to participate in app-based contact tracing. Using the largest patient-facing EHR in the NHS as an effective and timely questionnaire tool, we have revealed the role of uncertainties in both government messaging and not testing suspected COVID-19 infection in reducing willingness for app-based contact tracing and the importance of eliminating uncertainty in lockdown and virus-testing policies.

## Data Availability

The data described is held and controlled by Imperial College London NHS Foundation Trust.

## Appendix

### Contents

A. Week 7 of weekly longitudinal Care Information Exchange (CIE) Coronavirus Wellbeing Questionnaire for Patients **[attached as separate file]**
B. CONSORT flow diagram of response selection for inclusion in analysis
C. Map of UK showing registered CIE users by postcode.

## Appendix B.

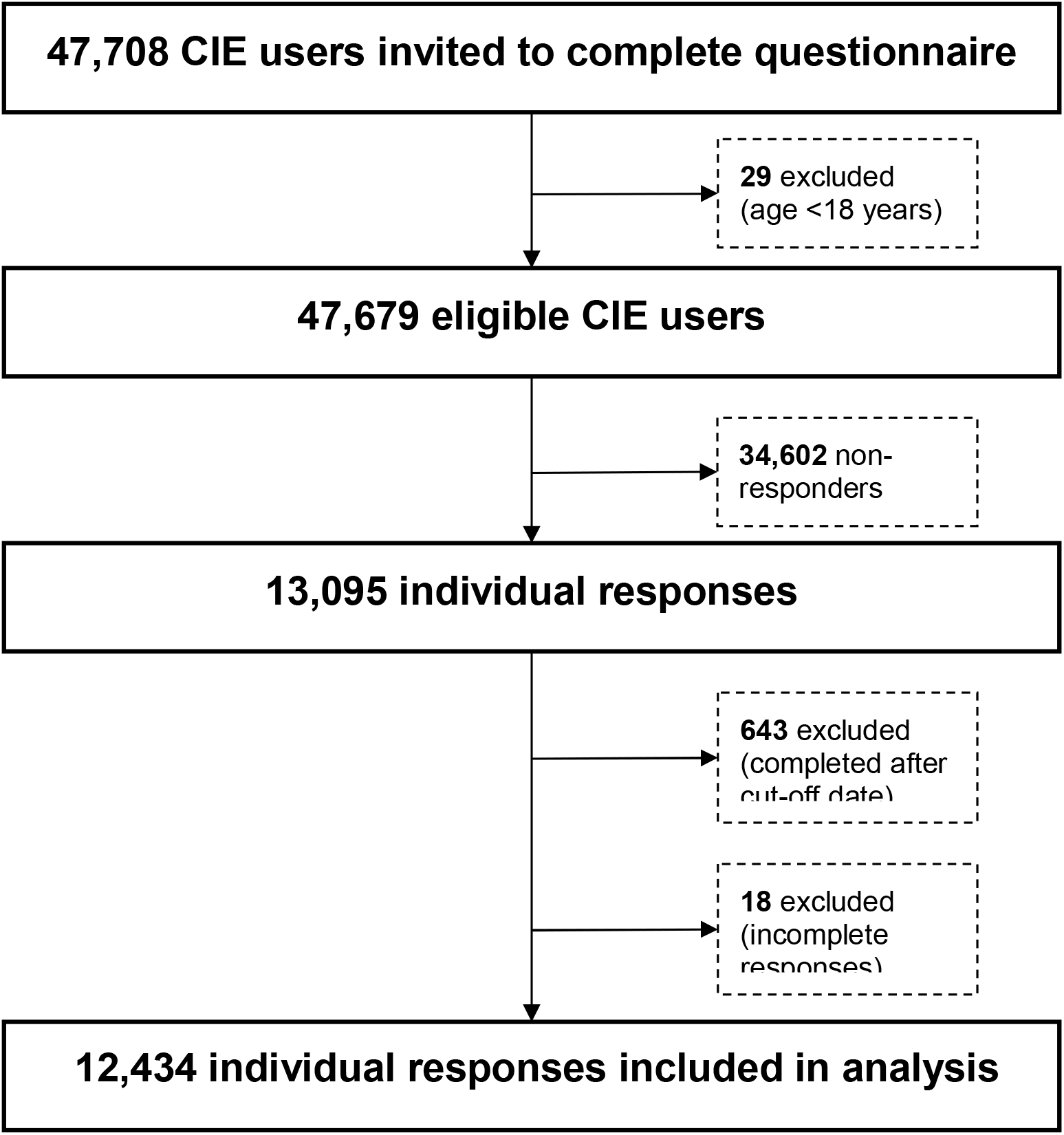

## Appendix C. Map of registered CIE users by postcode.

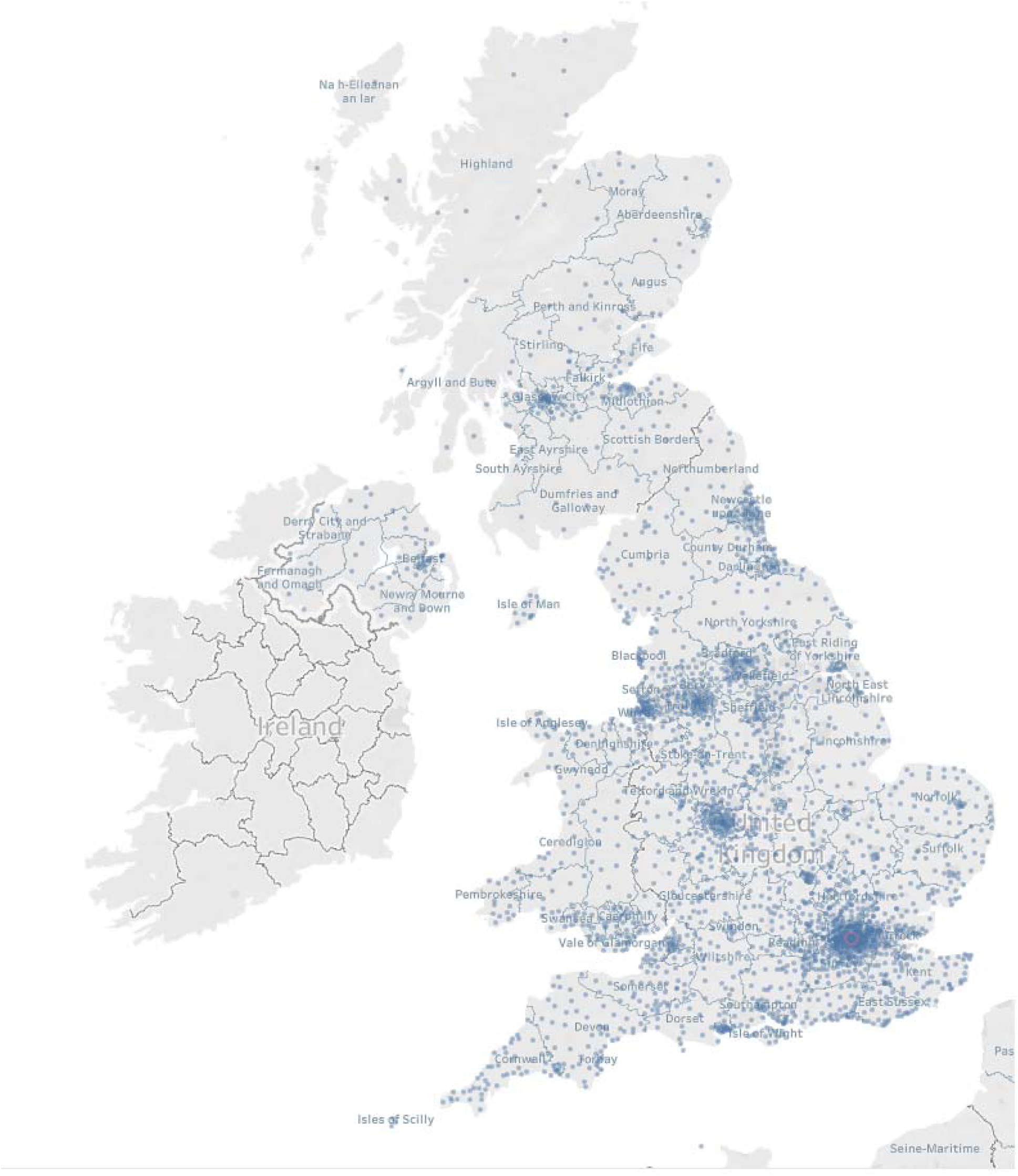

